# What keeps male amateur rugby union players committed to the game despite high risk of injury? A qualitative study

**DOI:** 10.1101/2024.08.18.24312177

**Authors:** Samuel Jake Lane, Colin Ayre

**Affiliations:** University of Bradford, Richmond Rd, Bradford, West Yorkshire, England, BD7 1DP; Bradford Teaching Hospitals NHS Foundation Trust, Duckworth Lane, Bradford, England BD9 6RJ

**Author notes:** **<u>Corresponding Author:</u>** Samuel Jake Lane, Present Address: 115 Maidenhall, Highnam, Gloucester, GL2 8DJ.

**Keywords:** Motivation, Participation, Sport, Rugby Union

## Abstract

**Objectives:** Rugby Union is a popular sport worldwide characterised by high intensity full contact match play. Injuries are common during matches and training. While musculoskeletal injury is expected in such a sport, there is emerging research evidence and increased public awareness of neurodegenerative disease risks. The objectives of the present study are to explore reasons for continued participation despite high risk of injury, gain understanding of perceived importance of participation and insight into why players may play with chronic injury.

**Design:** 10 male amateur rugby union players in Gloucestershire and Yorkshire aged 30±5.75 with 20±6.93 years of playing experience participated in semi-structured interviews with subsequent thematic analysis of transcripts.

**Results:** 4 major themes of identity, community, risk acceptance, and uniqueness of rugby were identified. None of the players reported plans to reduce participation in rugby.

**Conclusions:** Despite risks of injury there were no plans to reduce participation amongst players interviewed. Benefits gained from rugby are abundant and meaningful to players. Understanding these beliefs is helpful for medical staff supporting players, especially through injury. Overall, commitment to amateur rugby union remains strong despite new knowledge of the long-term risks involved. The findings show that amateur rugby union remains a popular sport with benefits extending beyond physical activity.

**Highlights:**

- There are 4 key themes regarding commitment to participation in male amateur rugby union despite risk of injury, regardless of new insights of potential long term injury risk and available injury epidemiology data
- Current players remain firmly committed to playing citing more benefits than negatives.
- Knowledge of risks is limited in the playing population recruited. This is significant as it highlights an issue that needs to be addressed regarding players ability to make an informed choice about their involvement in the game.

## Introduction

Rugby union is a contact sport with a high risk of injury during training and matches [1]. Due to the aggressive nature of the sport, musculoskeletal injuries are commonplace. Research into the epidemiology of rugby injuries has highlighted the high severity (days lost or matches missed) and incidence of injuries across amateur and professional populations, reporting the most common injury as concussion, with tackling being the most injurious action [2-6].

Former players are more likely to report suffering post-retirement effects of injury than former non-contact athletes [7] and increased difficulty with pain, mobility, and self-care compared to general population [8]. At the extreme, life changing permanent disability and even deaths have resulted from injuries sustained playing rugby union [9-11]

There is emergent evidence of increased risk of neurodegenerative disease associated with head contact from rugby collisions [12]. While this research was conducted on former professionals, the commonality of concussion across playing populations suggests this risk is apparent for amateurs.

Most existing literature examining motivation for participation has been conducted on female athletes and concerned factors for playing in the first instance, with an interest in gender roles and inclusivity [13-15]. However, key themes from those studies on female athletes including culture, sense of challenge, ‘love of the game’, and achievement were echoed in Dong et al’s [16] research on middle-aged male players including: making friends, the love of rugby (despite the risk and injuries), and self-actualisation. Aggression and physicality appear to be fundamental motivators with many interviewees highlighting these elements as the reason they were first attracted to or continue to play [13, 14, 16,17]. Madrigal et al [18] found passion for the sport and sport ethic to be major themes for playing through pain and injury in collegiate rugby, concluding that many will only stop with an external mandate (coach, parent, medical staff), or if impacting their activities of daily living. To the authors knowledge this is the first study investigating the reasons for participation in the amateur male playing population.

### Aims and objectives

There is a gap in the literature regarding the current male amateur rugby union playing population, specifically post emergent evidence of concussion risk and future health implications. It is hoped that an understanding of player beliefs and values may help develop empathy and improve patient centred care in rehabilitation. This may also shape discussions around medical retirement and how this can be done in a meaningful and supportive way, considering the players’ perceived importance of continued participation. There is also potential to provide context to why players are willing to play through injury or return to sport prematurely.

This study also aimed to determine if current players’ attitudes to injury risk and potential for long term health issues have been impacted by recent media coverage.

### Objectives

- To explore reasons for continued participation despite high risk of injury
- To understand the perceived importance of continued participation in rugby union
- To provide insight into reasons why players may continue to play with chronic injury.

## Methods

Ethical approval was granted by the Chair of the Biomedical, Natural, Physical and Health Sciences Research Ethics Panel at the University of Bradford on 12th October 2023 with amendments approved 13th November 2023. Convenience and snowball sampling techniques were utilised to recruit 10 current male amateur rugby union players over 18 years old in Gloucestershire and Yorkshire. All players involved were currently registered with amateur clubs at or below level 6 of the RFU pyramid. Participant information sheets were provided with written informed consent of participants obtained prior to involvement in the study. Audio recording and automated transcription of the interviews was done using Otter.ai. Transcriptions were corrected using original audio files, anonymised, and member checked by participants. Once final transcripts were confirmed audio recordings were destroyed. Interview transcripts were analysed using Braun and Clarke’s [19] reflexive thematic analysis method to deduce common themes to answer the research question.

## Results

Results of the main study produced major themes of *community, identity, uniqueness of rugby*, and *risk acceptance*. These were derived from the codes and initial themes seen in table 3. Of the 10 players interviewed 9/10 had suffered injuries as a direct result of playing rugby. Tables 1 and 2 highlight the nature and incidence of injuries sustained by the sample.

**Table 1.** Descriptive Statistics of Sample.

|  |  |
| --- | --- |
| **Age in Years (Mean±1SD)** | 30±5.75 |
| **Years Played (Mean±1SD)** | 20±6.93 |
| **Number of Injuries (Mean±1SD)** | 4.7±3.6 |
| **Nature of Injuries** | Fracture, Dislocation, Ligament Tear, Muscle Tear, Cartilage<br>Tear, Concussion, Nerve Damage |
| **Most Common Injury Site** | Knee 6/10, Hand 6/10 |

**Table 2.** Sample Injuries.

| Injured Area | # of Participants<br>Reporting Injury | Incidence |
| --- | --- | --- |
| Knee | 6 | 13 (MCL x5, ACL x3, Meniscus x4, Dislocation x1) |
| Hand/Fingers | 6 | 8 (Fracture x5, Dislocation x3) |
| Head/Face | 5 | 13 (Concussion x10, Nasal Fracture x3) |
| Shoulder | 4 | 5 (Fracture x3, Ligament x2) |
| Ankle | 4 | 3 (Ligament x2, Fracture x1) |
| Arm | 2 | 5 (Fracture x5) |
| Leg | 1 | 1 (Muscle Tear x1) |
| Nerve Damage | 1 | 1 (Nerve Damage x1) |

**Table 3.**
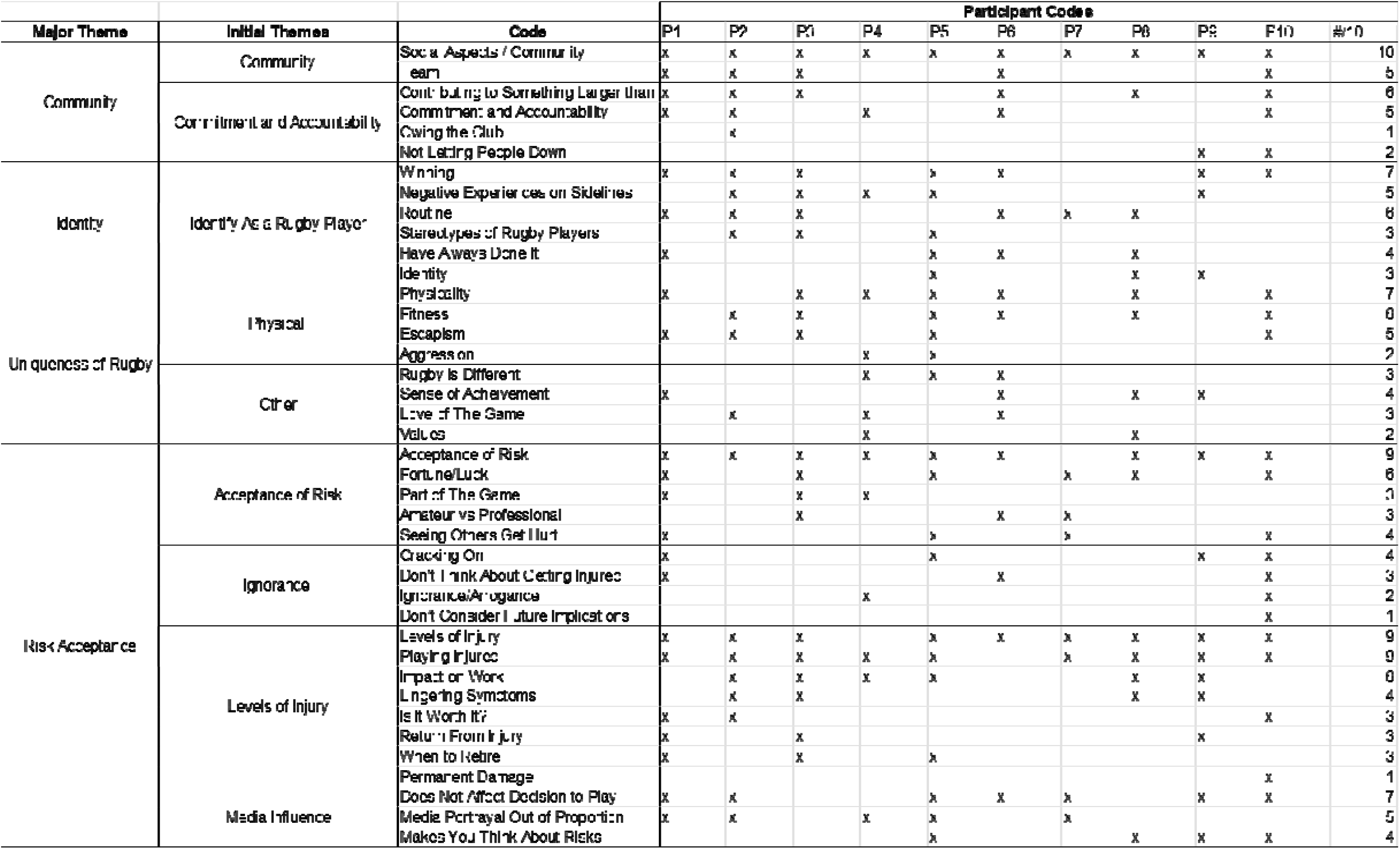
Thematic Analysis Breakdown.

### Community

All participants reported the community and social aspects of rugby as key reasons for remaining committed to the game, with 6/10 mentioning the contribution towards a collective goal or something bigger than themselves. There were numerous mentions of the club as a social hub on game days beyond. Friendships that were formed playing rugby also appeared to go much further than the 80 minutes of gameplay.

> **P3** *“So the biggest one is obviously the friendships. Many… well, all of my close friends play rugby with me… not only are we friends off the pitch socially, but it is a way of bonding together in a collective cause…”*
>
> **P6** *“I think there’s great joy to be found when you’re playing, you’re buying into a common goal… And then you come through it all together, win or lose and you’ve done it in a way that was difficult, tiring, painful…”*

### Identity

Winning and the feeling of elation having worked hard with others was a key element of continued participation. 6/10 players discussed rugby as being part of their weekly routine and would suffer without this.

Participants had played an average of 20±6.93 years and 4/10 went as far as saying they “had always done it”. The sense of identity as a rugby player was most evident when participants discussed negative experiences of being sidelined with injury and how they would struggle if they could no longer play.

> **P3** *“you feel like you’re not part of the team, you’re not part of squad and… I found that quite stressful and quite difficult*.*”*
>
> **P4** *“…you’re not really having an input apart from cheering them on…you’re still their mate…but you’re essentially just a fan*.*”*
>
> **P5** *“… I’d say it’s almost a bit like a badge of honour … and if I were to stop playing, would it feel like a bit of a fraud thing? A has-been sort of thing? I think that’s an important part of it psychologically… that probably is what keeps me still going secretly*.*”*
>
> **P10** *“I remember feeling… almost outcast… like I wasn’t kind of needed or it didn’t matter to anyone that I wasn’t playing or wasn’t there”*

### Uniqueness of Rugby

The uniqueness of rugby could be categorised as physical, mental, and other aspects. Players highlighted the physicality of contact sport and a fun way to maintain fitness as being a strong motivating factors for playing the game. Mental health and escapism from daily worries were discussed by 7/10 and 5/10 players respectively. Players struggled to define exactly what differentiates rugby from other sports. Common remarks were the excitement of the changing room, the feeling of winning, and the physical demands required for success.

> **P1** *“When you play the game you forget about everything else… you’re in that moment, and you’re focused on the task at hand and nothing else, you don’t think about anything else, I don’t find that I think about how stressful my week is. It actually all goes away”*
>
> **P3** *“…the certain type of person that wants to play rugby and enjoys that competitive aspect is… a physical person who enjoys contact and enjoys that confrontational element. So if you were to water down the game too much, I don’t think it would be rugby*.*”*
>
> **P10** *“ I’d miss the experience with your friends in the changing room especially before, the build-up, the excitement…”*

### Risk Acceptance

90% of players were prepared to accept the risks of playing a contact sport for the benefits they feel they gain with 7/10 stating that the risks presented in the media do not impact their decision to continue to play. Injuries were seen as part of the game, and 5/10 felt that the media portrayal of risks was potentially out of proportion.

9/10 players had played through injury although the level of injury was a large factor in whether players continued to play. A major factor in deciding to continue was the impact or potential impact on work. 6/10 players felt they had been lucky in their careers based on the number and severity of the injuries they had suffered.

> **P4** *“…it’s part and parcel of it really. If you and go into it and are worrying about an injury my advice is don’t play*.*”*
>
> **P5** *“ I would consider myself relatively lucky in terms of the injuries that I’ve sustained because they haven’t been you know, life altering or career ending*.*”*
>
> **P7** *“it’s not impacted my personal perception of playing. For me I think the risks involved possibly been over exaggerated. So I think… you kind of know what you’re getting into…”*

## Discussion

To the author’s knowledge this is the first study into reasons for continued participation in amateur rugby union despite high risk of injury and emerging evidence of neurodegenerative disease risk. Analysis of interviews found 4 key themes of *Community, Identity*, Risk Acceptance, and the *Uniqueness of Rugby* to be the major drivers commitment. An appreciation and understanding of the risks involved in continued participation is apparent, however contemplation of behaviour change (quitting the sport) in the population recruited is not evident. Rather, many strong positives associated with participation were found which outweigh these risks for players.

Media reporting of emergent evidence and exposure to former players suffering neurodegenerative disease secondary to concussion appears to have positively impacted the handling of concussion injuries and player consideration of playing through injury.

### Community

Community and social aspects of the sport were major factors for continued participation for all participants. One major finding was that social aspects of the club environment were mentioned more than experiences of physically playing the sport. 6/10 participants described the contribution to something larger than themselves as a positive aspect of participation. This act of service and contribution has been linked to a sense of meaning in life and is associated with greater happiness and wellbeing [20].Community and the associated friendships and commitment to others has been reported by existing research into rugby participation[13,16]. However, those studies did not consider the population of comparatively young males engaging in competitive amateur sport used in the present study – arguably the largest stakeholder population of the sport.

### Identity

Identifying as a rugby player appears to be important to participants and potentially explains justification for continuing to play even after suffering injury, with 90% of players having returned to the sport after multiple injuries. Players also discussed long term involvement in the sport with 4/10 stating they have always done it and 6/10 regarding training and matches as a large part of their weekly routine.

The sport ethic is a term used to describe the behaviours of sacrifice, risk taking, seeking distinction, and pushing limits evident in competitive sportspeople [21] and referenced as a contributing factor for playing through pain and injury in collegiate rugby players [18]. While the sport ethic was not mentioned explicitly by any participants, these values and behaviours were evident and should be recognised by medical professionals who work within rugby union as a reason for playing through injury or premature desire return to competition. 5/10 participants reported negative experiences of being sidelined with injury and perceived loss of player identity. This is consistent with the findings of Murphy and Sheehan [22] who found the psychological burden of injured players can negatively impact recovery and outcome of rehabilitation. This suggests a sense of belonging that is lost when the player is sidelined, with the physical act of playing the sport being a prerequisite to being a part of the team. This finding is of particular interest to health professionals supporting injured players who should endeavour to include the player in the team dynamic in whatever way they can, for example in water carrier or coaching roles during training and matches that they are unable to play in.

### Risk Acceptance

All players understood the musculoskeletal risk associated with playing a contact sport. They accept this risk and continue playing due to the perceived benefits of participation. Players believed they and other players “knew what they were getting into” yet paradoxically had little to no knowledge of the emergent evidence of the risk of neurodegenerative disease associated with rugby collisions beyond media headlines.

Media portrayal of rugby and the associated risks was overall seen in a negative light. Players discussed the sensationalist nature of the media, yet 9/10 players had suffered substantial physical injury with 1 recalling difficulties with light sensitivity and vomiting after concussion. There appears to be some conflict between reported experiences, publicly available epidemiology figures, and player perceptions of the risks involved. This cognitive dissonance has been seen previously in rugby literature concerning parent perceptions of risk in youth contact rugby [23]. This suggests perhaps an element of denial from players and that sources disseminating information regarding risks needs to come from a source regarded as trustworthy.

While at the top flight, clubs have extensive medical departments, in regional rugby, only 23% of clubs were found to be compliant with RFU regulations regarding medical personnel, facility, and equipment provision and 30% of clubs were unable to confirm medical personnel had required qualifications [24]. With such poor compliance to player welfare regulations below the top level, the decision to accept the risks of injury and play is ultimately on the player. However, it remains important that players have access to trustworthy information in order to make informed decisions regarding participation. Based on the findings of this study this remains an area for improvement.

### Uniqueness of Rugby

The sanctioned aggression, physically demanding contribution to a shared goal, and social aspects that extend beyond the time on the field appear to be elements of the sport that players are unable to experience in other sports that they participate in e.g. football and golf. Factors such as the contact element (7/10) and the collective work towards something bigger than the self (6/10) would be missed should players end participation. Senecal [25] discussed the difficult transition faced by contact athletes at the end of their sporting careers and the difficulty in replacing the ‘peak experiences’ felt when playing. The themes of alteration in time, magnitude of joy and elation, and absorbed awareness found in that study are consistent with the aspects of rugby that participants in the current study reported. These findings, along with the reported negative experiences of being unable to participate while sidelined, suggest that it may be pertinent for players to consider life after rugby and be prepared for such a transition. This is something that sets rugby apart from other sports. Although veterans and touch or walking versions of the game are available, participation in the full contact, open age iteration has a time limit due to its physically demanding nature.

### Unexpected Results

None of the participants expressed planning to reduce participation in rugby suggesting that the sample used are in the pre-contemplative phase of behaviour change [26]. Half (5/10) of the participants had a defensive stance against the media narrative of rugby being dangerous. It is possible that this is unique to amateur players at this level who potentially do not undergo the education that professional players may have access to via professional medical staff. Another potential reason for this could be wilful ignorance and a desire to avoid an uncomfortable truth. This is comparable to smokers who understand the risks and may suffer because of their actions but continue anyway [27]. Likewise with smoking addiction and the associated neurochemical change, rugby and the benefits derived potentially have ‘addictive’ characteristics. While exercise addiction is well documented [28] it is beyond the scope of this study to suggest this was evident in the players interviewed. It is, however, well known that exercise has significant effects on the neurochemistry of the brain [29] and the loss of belonging, stress, and low mood associated with being sidelined through injury parallels withdrawal symptoms.

## Conclusions

Player’s actions and behaviours toward participation remain unchanged despite the understanding, observation, and lived experience of musculoskeletal injury risk. Emergent evidence of neurodegenerative disease risks appeared to be not fully appreciated or understood (or potentially disregarded). The benefits that players gain from their involvement in rugby are both abundant and meaningful to them and can be organised into the major themes of *identity* and *community*, while the themes of *risk acceptance* and *uniqueness of rugby* provide insight into the justification of the choice to continue to play the sport.

Whilst these results cannot be generalised, the present study shows current players have a continued desire to be involved and that perceived benefits strongly outweigh the perceived risks of participation. It is uncertain whether further information or deeper understanding of risk would change this, though players do not appear to be contemplating behavioural change.

### Limitations

A small sample size (10) from 2 geographical locations (Gloucestershire and Yorkshire) reduces generalisability of results, therefore future research should consider larger samples from more varied populations. Use of a solo researcher prevented triangulation of qualitative analysis coding and theme development. The researcher is a former rugby player and therefore may have some inherent personal biases. Interviewees may have felt an inclination to conform due to conversing with a former rugby player, however individual interviewing and anonymised transcription allowed for open and honest responses. Total injuries were difficult to accurately report as many players could not give precise injury histories due to memory and the difficulty in knowing what would constitute an injury. Ex-players or players that have sustained injury and were not intending to return to play are likely to have provided different responses to the sample selected, however this study concerned reasons for remaining committed as opposed to consensus of players (current or former) on injury risk. This is an area that could be explored in future to compare against results of the present study.

### Recommendations

The beliefs and actions of players is apparently contradictory with many players stating that they “know what they are getting into” playing the game, while simultaneously appearing to have limited knowledge of the neurodegenerative risks involved. This suggests efforts to educate players about these risks have been unsuccessful or that players are ignorant and avoidant of the data presented to them.

Non-biased education delivered by medical professionals or via governing bodies such as the RFU incorporating current or former players may aid in greater acceptance by leaning into the themes of identity and community.

## Data Availability

All data produced in the present study are available upon reasonable request to the authors

## Appendices Appendix A. Interview Schedule

### Interview Schedule

#### Schedule items

1. What positives do you feel you gain from playing rugby?
2. What are your experiences with injury and does that factor into continued participation?
3. Have you continued to participate with symptoms or lasting effects from previous injuries?
4. What would you miss if you stopped playing?
5. What keeps you committed to the sport?
6. How do you feel about the recent media portrayal of risks involved and has this impacted your perception on playing?

## Ethical Approval

Ethical approval was granted by the Chair of the Biomedical, Natural, Physical and Health Sciences Research Ethics Panel at the University of Bradford on 12th October 2023 with amendments approved 13th November 2023.

